# BNT162b2 mRNA Covid-19 vaccine does not impair sperm parameters

**DOI:** 10.1101/2021.04.30.21255690

**Authors:** Myriam Safrai, Benjamin Reubinoff, Assaf Ben-Meir

**Affiliations:** In Vitro Fertilization Unit, Hadassah Medical Organization and Faculty of Medicine, Hebrew University of Jerusalem, Israel

## Abstract

Mass vaccination using newly approved vaccines against the severe acute respiratory syndrome coronavirus 2 (SARS-CoV-2) has begun globally. However, their effect on fertility have not yet been investigated. Previous studies demonstrate that SARS-CoV-2 infection may impair sperm parameters. In this study, we are the first to assess the effect of the BNT162b2 mRNA Covid-19 vaccine on sperm parameters. Our results demonstrate that the vaccine does not impair sperm parameters. Thus, we recommend that couples desiring to conceive should vaccinate, as vaccination does not affect sperm whereas SARS-CoV-2 infection does impair sperm.

## Introduction

Mass vaccination using newly approved vaccines against the severe acute respiratory syndrome coronavirus 2 (SARS-CoV-2) has begun globally. To eradicate this disease, vaccination of the younger population is recommended (1). A number of clinical trials have been done to assess the safety of these vaccines, however, their effect on fertility have not yet been investigated (2). Nevertheless, the Task Force does not recommend withholding vaccines from patients who are planning to conceive (3). Furthermore, the gonads may potentially be vulnerable to SARS-CoV-2 infection (4), and several studies found a significantly negative impact of SARS-CoV-2 infection on sperm parameters (5).

## Material and methods

To assess the effect of the BNT162b2 mRNA Covid-19 vaccine on male fertility, we collected data from all patients in the IVF units of a large university hospital between February and March 2021, after vaccination of the population began. The study was approved by the institutional review board (IRB-0092-21-HMO). A total of 43 male patients undergoing IVF in our service (14 due to male factor, and 29 with normal spermogram results), had been vaccinated. Medical records of these patients prior to vaccination were retrospectively reviewed (PRE vaccination) using the hospital’s electronic database, and was compared prospectively to collected data of all patients after BNT162b2 vaccine (POST vaccination). To minimize bias, each patient served as a self-control before and after vaccination. Parameters are presented as mean ± standard deviation. Comparisons between pre- and post-vaccination measurements were conducted with paired t-tests. A *p*-value of ≤0.05 was considered significant.

## Results

The clinical characteristics and spermogram parameters are shown in Table 1. Sub-group analyses were performed for patients with male infertility, and patients with normal spermogram results undergoing IVF for indications other than male infertility. None of the parameters differed significantly after vaccination.

**Table 1.** Patient characteristics and spermogram results before and after BNT162b2 vaccination.

|  |  |  |  |
| --- | --- | --- | --- |
| Age (years) | 37.1 ±6.6 |  |  |
| Time from the first vaccine dose (days) | 33.6 ± 20.2 |  |  |
|  | PRE vaccination | POST vaccination | *p*-value |
| **All patients (n=43)** |  |  |  |
| Sperm volume (ml) | 3.0 ± 1.5 | 2.9 ±1.7 | 0.8 |
| Sperm concentration (10<sup>6</sup>/ml) | 43.6 ±58.0 | 47 ±54.8 | 0.7 |
| TMC (10<sup>6</sup>) | 48.5 ±83.4 | 61.7 ±92.9 | 0.4 |
| **Patients with normosperm (n=29)** |  |  |  |
| Sperm volume (ml) | 3.2 ±1.6 | 3.2 ±1.8 | 0.4 |
| Sperm concentration (10<sup>6</sup>/ml) | 60.9 ±62.4 | 67.8 ±56.4 | 0.6 |
| TMC (10<sup>6</sup>) | 71 ±93.9 | 89.9 ±102.4 | 0.5 |
| **Patients with male infertility (n=14)** |  |  |  |
| Sperm volume (ml) | 2.6 ±1.1 | 2.3 ± 1.3 | 0.4 |
| Sperm concentration (10<sup>6</sup>/ml) | 4.8 ±7.7 | 8.4 ±17.4 | 0.4 |
| TMC (10<sup>6</sup>) | 1.9 ±4.1 | 6.2 ±14.4 | 0.2 |

All continuous variables are expressed as mean ± standard deviation (SD); TMC=Total motile count

## Conclusion

Our study is the first to evaluate the impact of the BNT162b2 vaccine on sperm parameters. Each patient serving as its own control increased accuracy and demonstrated that this vaccine appeared to have no impact on sperm parameters. These preliminary results are reassuring to the young male population undergoing vaccination worldwide. Given that SARS-CoV-2 infection may impair male fertility (4, 5), couples desiring to conceive should vaccinate, as vaccination does not affect sperm, whereas SARS-CoV-2 infection does impair sperm parameter.

## Data Availability

The data that support the findings of this study are available on request from the corresponding author, [MS].

## References

1. Dagan N, Barda N, Kepten E, et al. BNT162b2 mRNA Covid-19 vaccine in a nationwide mass vaccination setting. N Engl J Med Feb 24, 2021.

2. Coronavirus/COVID-19 Task Force of the American Society for Reproductive Medicine. American Society for Reproductive Medicine (ASRM) Patient Management and Clinical Recommendations during the Coronavirus (COVID-19) Pandemic: UPDATE No. 11 – COVID-19 Vaccination. [Internet]. 2020 [cited 2021 Mar 20]; Available from: https://www.asrm.org/globalassets/asrm/asrm-content/news-and-publications/covid-19/covidtaskforceupdate11.pdf?utm_source=Informz&utm_medium=email&utm_campaign =EmailBlast

3. Khalili MA, Leisegang K, Majzoub A, et al. Male fertility and the COVID-19 pandemic: systematic review of the literature. World J Mens Health 2020;38(4):506–520.

4. Patel DP, Punjani N, Guo J, Alukal JP, Li PS, Hotaling JM. The impact of SARS-CoV-2 and COVID-19 on Male Reproduction and Men’s Health. Fertil Steril 2021; S0015-0282(20)32780-1.

5. Gacci M, Coppi M, Baldi E, Sebastianelli A, Zaccaro C, Morselli S, Pecoraro A, et al. Semen impairment and occurrence of SARS-CoV-2 virus in semen after recovery from COVID-19. Hum Reprod 2021.

